# At-Home High-Intensity Interval Training for Individuals with Paraplegia Following Spinal Cord Injury: *A Pilot Study*

**DOI:** 10.1101/2023.06.21.23291711

**Authors:** Margaux B. Linde, Kevin L. Webb, Daniel D. Veith, Olaf H. Morkeberg, Megan L Gill, Meegan G. Van Straaten, Edward R. Laskowski, Michael J. Joyner, Lisa A. Beck, Kristin D. Zhao, Chad C. Wiggins, Kristin L. Garlanger

**Affiliations:** Mayo Clinic, Rehabilitation Medicine Research Center, Department of Physical Medicine & Rehabilitation, Rochester, MN; Mayo Clinic, Department of Anesthesiology & Perioperative Medicine, Rochester, MN; Mayo Clinic, Department of Physical Medicine & Rehabilitation and Division of Sports Medicine, Department of Orthopedics, Rochester, MN

**Keywords:** Aerobic training, Cardiovascular health, Rehabilitative medicine, and Arm ergometry

## Abstract

**Objective:** This pilot study aimed to assess the efficacy of a 16-week at-home high-intensity interval training (HIIT) program among individuals with spinal cord injury (SCI).

**Methods:** Eight individuals (age: 47±11 (SD) years, 3 females) with SCI below the sixth thoracic vertebrae participated in a 16-week at-home HIIT program using an arm ergometer. Participants completed baseline graded exercise tests to determine target heart rate zones. HIIT was prescribed thrice per week. Each training session consisted of six one-minute bouts with a target heart rate ∼80% heart rate reserve (HRR), interspersed with two minutes of recovery at ∼30% HRR. A portable heart rate monitor and phone application provided visual feedback during training and allowed for measurements of adherence and compliance. Graded exercise tests were completed after 8 and 16 weeks of HIIT. Surveys were administered to assess participation, self-efficacy, and satisfaction.

**Results:** Participants demonstrated a decrease in submaximal cardiac output (*P*=0.028) and an increase in exercise capacity (peak power output, *P*=0.027) following HIIT, indicative of improved exercise economy and maximal work capacity. An 87% adherence rate was achieved during the HIIT program. Participants reached a high intensity of 70% HRR or greater during ∼80% of intervals. The recovery HRR target was reached during only ∼35% of intervals. Self-reported metrics of satisfaction and self-efficacy with at-home HIIT scored moderate to high.

**Conclusion:** Participants demonstrated an improvement in exercise economy and maximal work capacity following at-home HIIT. Additionally, participant adherence, compliance, satisfaction, and self-efficacy metrics suggest that at-home HIIT was easily implemented and enjoyable.

## INTRODUCTION

Individuals with spinal cord injury (SCI) often experience motor impairments below the level of injury, leading to worsened levels of physical activity, a greater prevalence of cardiovascular disease, and reduced life expectancy (1). For instance, some evidence has suggested that the prevalence of heart disease may be ∼3 times greater among individuals with SCI compared to able-bodied individuals (17.1% versus 4.9%, respectively, (2)). Regular exercise is critical for mitigating cardiometabolic disease risk factors such as hypertension, dyslipidemia, glucose intolerance, and obesity (3–7). Current guidelines recommend ∼90 minutes or more of moderate to vigorous aerobic exercise per week to improve cardiometabolic outcomes among individuals with SCI (8). However, those with SCI experience substantial barriers to exercise, such as poor access to facilities, unaffordable equipment, and fear of self-injury (9).

High-intensity interval training (HIIT) is a method of exercise where individuals engage in repeated intervals of intense activity, each followed by a brief period of recovery. HIIT has previously demonstrated substantial improvements in cardiometabolic health in the able-bodied population (10–12). Additionally, HIIT may shorten the duration of activity required to elicit similar cardiometabolic outcomes compared to continuous, moderate-intensity aerobic exercise (13,14). Shortening the activity duration may mitigate repeated-use injuries in the upper extremities for individuals with SCI and improve adherence to a prescribed exercise routine. For these reasons, recent research has investigated the utility of HIIT as a promising method to improve cardiometabolic health among individuals with SCI. However, current research is limited to examining HIIT with training sessions located in a laboratory setting rather than at home, hindering the generalizability of these findings (15–17).

As such, this pilot study aimed to assess the efficacy of a 16-week at-home HIIT program among individuals with paraplegia due to SCI. Overall, we hypothesized that participants would demonstrate an increase in physical fitness (i.e., improvement in peak oxygen uptake (VLJO_2_peak) and peak power output) and have high training adherence and compliance.

## MATERIALS AND METHODS

### Study Design

All experimental procedures were approved by the Mayo Clinic Institutional Review Board (IRB# 18-004972, Clinical Trials Identifier: NCT04378218). Experimental procedures were performed in accordance with the ethical standards set by the *Declaration of Helsinki*, excluding registration in a public database. Participants provided written informed consent prior to enrollment.

### Participants

Ten participants diagnosed with SCI below the sixth thoracic vertebrae (T6) were enrolled in this clinical trial. Following enrollment, one participant chose to withdraw from the trial. Another participant completed the trial but was removed from the dataset due to discovery of a cervical (C7) lesion level (finding occurred during subsequent screening for another study, contradictory to medical record indicating a T7 lesion level). Thus, eight eligible participants completed the trial, presented in **Table 1**. To be eligible for inclusion, participants must have been at least 18 years of age and reported the use of a manual wheelchair as a primary means of mobility. Individuals were excluded if the SCI occurred at or above T6 due to confounding influences on cardiac autonomic innervation (18). Additionally, individuals were excluded if the injury occurred less than six months prior to enrolment or if they were diagnosed with any contraindicated health condition for participation in an exercise program. None of the participants were taking blood pressure medications during the study.

**Table 1.**
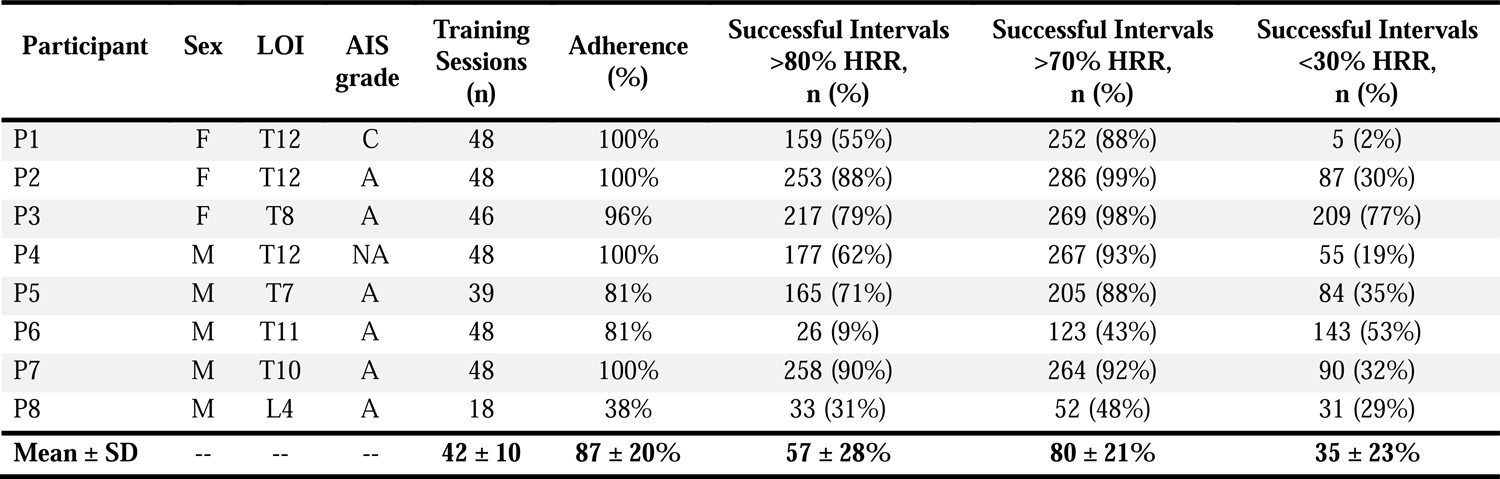

| Participant | Sex | LOI | AIS grade | Training Sessions (n) | Adherence (%) | Successful Intervals >80% HRR, n (%) | Successful Intervals >70% HRR, n (%) | Successful Intervals <30% HRR, n (%) |
| --- | --- | --- | --- | --- | --- | --- | --- | --- |
| P1 | F | T12 | C | 48 | 100% | 159 (55%) | 252 (88%) | 5 (2%) |
| P2 | F | T12 | A | 48 | 100% | 253 (88%) | 286 (99%) | 87 (30%) |
| P3 | F | T8 | A | 46 | 96% | 217 (79%) | 269 (98%) | 209 (77%) |
| P4 | M | T12 | NA | 48 | 100% | 177 (62%) | 267 (93%) | 55 (19%) |
| P5 | M | T7 | A | 39 | 81% | 165 (71%) | 205 (88%) | 84 (35%) |
| P6 | M | T11 | A | 48 | 81% | 26 (9%) | 123 (43%) | 143 (53%) |
| P7 | M | T10 | A | 48 | 100% | 258 (90%) | 264 (92%) | 90 (32%) |
| P8 | M | L4 | A | 18 | 38% | 33 (31%) | 52 (48%) | 31 (29%) |
| **Mean ± SD** | -- | -- | -- | **42 ± 10** | **87 ± 20%** | **57 ± 28%** | **80 ± 21%** | **35 ± 23%** |

### Laboratory Testing

Participants attended three laboratory testing sessions during the study: 1) at baseline, 2) after eight weeks of HIIT, and 3) after 16 weeks of HIIT. Due to COVID-19 related institutional closures, graded exercise tests performed at baseline, week 8, and week 16 of HIIT were only available in five participants. Each laboratory visit included a graded exercise test to task failure using an arm cycle ergometer (Model 891E Upper Body Ergometer, Monark Exercise AB, Vansbro, Sweden). Following familiarization with the test protocol and arm cycle, participants selected a comfortable pedaling cadence between 60 and 70 revolutions per minute and were instructed to maintain this cadence during the test within three revolutions per minute. Because of the mechanical nature of the arm cycle, starting workloads varied for each participant due to the individually selected cadence. Each graded exercise test began with two minutes of unloaded pedaling followed by gradual increases in resistance by adding 0.1-0.2 kg to the flywheel every two minutes until task failure. Starting power output and stepwise increases for all participants was 15±3 W (Range 12-20 W). The graded exercise test protocols (cadence and stepwise increases in power output) were held similar for each participant between laboratory visits. Resting and peak heart rate obtained from the baseline graded exercise test (before HIIT) were used to determine individualized target heart rates for at-home training sessions.

For each graded exercise test, participants breathed through a mouthpiece connected to a three-way T-shape non-rebreathing pneumatic sliding valve (Series 8500, Hans Rudolph, Shawnee, KS, USA) and were instrumented with a 12-lead ECG and patient monitoring system (Cardiocap/5, Datex, Louisville, CO, USA). During the graded exercise tests, breath-by-breath respiratory measurements were performed using a cardiorespiratory diagnostic system (MGC Diagnostics, St. Paul, MN, USA) interfaced with a mass spectrometer (MGA-11000, Perkin Elmer, Waltham, MA, USA). Respiratory gas exchange measurements and heart rate were averaged across the last 30 seconds of each stage of the graded exercise test. VLJO_2_peak was taken as the average of breaths during the last 30 seconds prior to task failure.

Cardiac output was measured during the graded exercise tests using a previously described open-circuit acetylene wash-in technique (19,20). Cardiac output was assessed three times during each graded exercise test; 1) at rest, 2) during exercise at a submaximal intensity (respiratory exchange ratio ∼1.0 during baseline visit), and 3) immediately prior to task failure. Briefly, participants were switched from inspiring room air to a pre-mixed, commercially available wash-in gas mixture stored in a Douglas bag containing 0.9% helium, 0.6% acetylene, 21% oxygen, and balanced with nitrogen. Upon switching, participants were instructed to breathe normally for 8-12 breaths. Cardiac output was then estimated from the rate of disappearance of acetylene. Stroke volume was taken as the quotient of cardiac output and heart rate.

### At-Home HIIT Training

Each participant was instructed to complete three at-home HIIT training sessions per week, totaling 48 structured HIIT sessions over 16 weeks. Each training session was 24 minutes in length and structured as follows: three-minute warmup, six one-minute high-intensity intervals with a target intensity at 80% of the individuals’ heart rate reserve (HRR, calculated using the Karvonen formula) with two minutes of active recovery, and a three-minute cooldown (**Fig 1A and 1B**). Heart rate targets were kept consistent throughout the training period. Of note, the heart rate kinetics during the first one to two intervals were often not rapid enough to achieve the target heart rate of 80% HRR. Therefore, if a participant reached a threshold of >70% HRR, we deemed the interval acceptable in achieving the high-intensity target.

**Figure 1.**
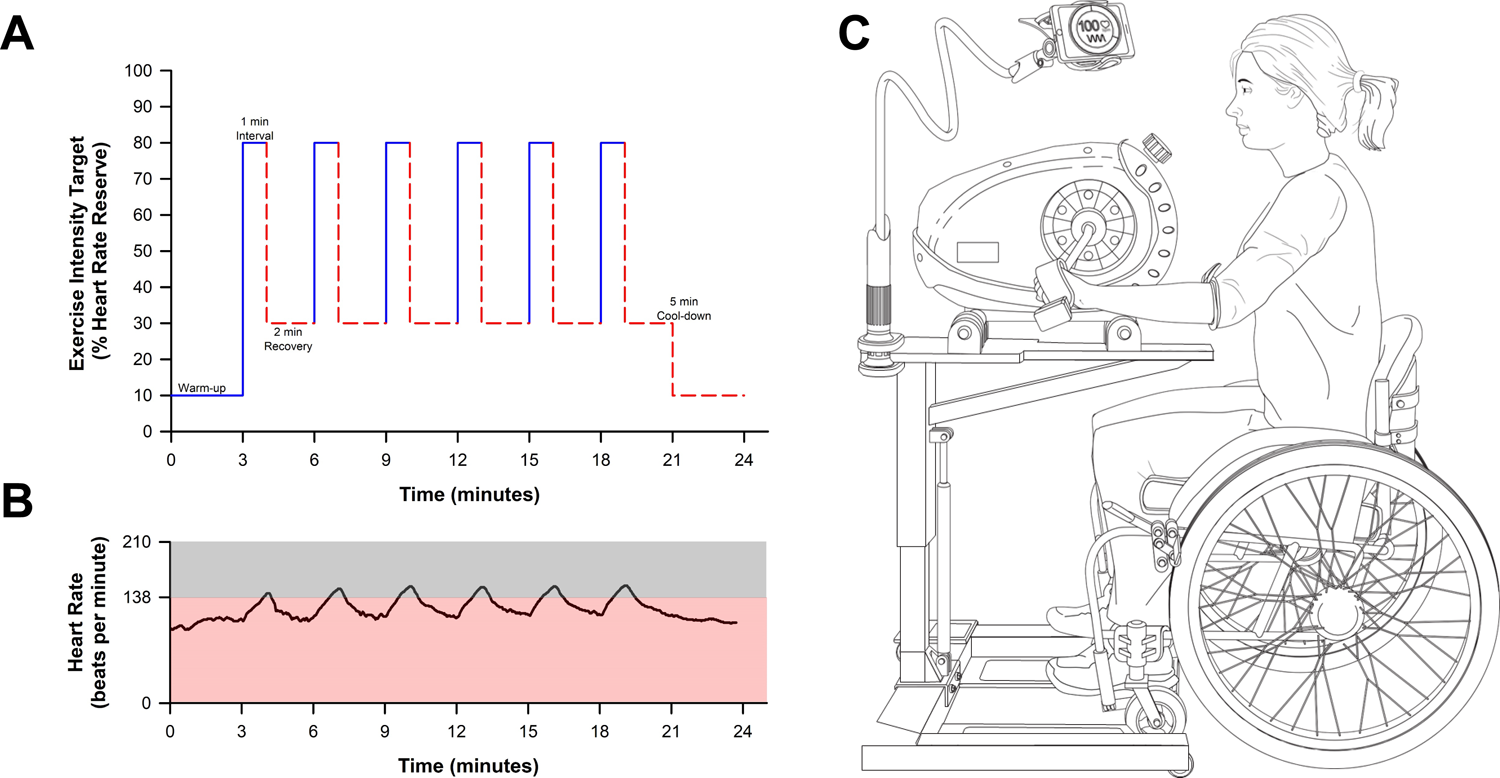
At-home high-intensity interval training (HIIT) study design. **A)** Schematic depicting an example of an at-home HIIT session. The target exercise intensity (% of heart rate reserve) is depicted for one-minute bouts of high intensity followed by two minutes of recovery. **B)** Representative figure of participant heart rate data during an at-home HIIT session obtained from the Polar phone application. The shaded grey-area depicts the target heart rate zone corresponding to ∼80% HRR. **C)** Schematic of the participant equipment provided for at-home HIIT. Participants were given an arm cycle, hydraulic table, and Polar heart rate strap to provide visual feed-back and instruction to achieve target exercise intensity during the training sessions. The participant’s phone was mounted to the hydraulic table to allow for visual heart rate feedback.

All at-home training sessions were performed using an arm cycle mounted to a wheelchair-accessible hydraulic table (PhysioTrainer UBE and Hydraulic UBE Table, HCI Fitness, Langley, WA, USA). Each in-home unit was provided and installed in the participants’ homes by the study team (**Fig 1C**). Each participant was provided with a portable heart rate monitor (Polar H10 Heart Rate Sensor, Polar Electro Inc., Bethpage, NY, USA) that was integrated with a smartphone using a commercially available applications (Polar Beat and Polar Coach Applications, Polar Electro Inc., Bethpage, NY, USA). Audio recordings, created by the study team and Mayo Clinic Media Services Department, were available to provide motivation to achieve prescribed intensity and interval timing. Sessions were guided by real-time visual heart rate through the phone application. Heart rate and interval durations during the training sessions were measured using the phone application. From these data, participant adherence (completion of prescribed training sessions) and compliance (the achievement of target heart rate zones during bouts of high intensity and recovery) were determined.

### Survey Assessments

Several surveys were developed for this study to determine participant-reported levels of safety (**Supplementary Table 1**). Phone calls were initiated when surveys went unreturned to ensure accurate tracking of adverse events. Participant communication excluded any coaching or encouragement to avoid confounding investigator influence. Additionally, the SCI Exercise Self-Efficacy Scale (SCI ESES) was used to assess participants’ confidence regarding carrying out regular exercise (21). The SCI ESES consists of a 4-point rating scale (1: not at all true, 4: always true). The SCI ESES was completed at three time points (before, during, and after HIIT) to assess changes in confidence in performing exercise. A final survey, administered after the study, was developed to assess participant satisfaction with the at-home HIIT exercise program (**Table 2**).

**Table 2.**

| Participant | Q1 | Q2 | Q3 | Q4 |
| --- | --- | --- | --- | --- |
| P1 | Yes | Yes | Yes | Yes |
| P2 | No | Yes | Yes | No |
| P3 | Yes | Yes | Yes | No |
| P5 | Yes | Yes | Yes | No |
| P6 | Yes | Yes | Yes | No |
| P7 | Yes | Yes | Yes | Yes |
| P8 | Yes | Yes | No | Yes |
| **Survey Questions:**<br>Q1: Do you like performing this HIIT exercise program?<br>Q2: Would you recommend HIIT exercise to a friend?<br>Q3: Would you continue performing HIIT as part of your exercise routine?<br>Q4: Did your physical activity increase outside of the HIIT exercise program?<br>Satisfaction survey questions and results for each participant following 16 weeks of at-home HIIT. Note, participant four (P4) did not complete the survey. Abbreviations: Q, question. |  |  |  |  |

### Data Analysis

Normality of data were confirmed using Shapiro-Wilk tests. Physiological data (heart rate, cardiac output, stroke volume, VLJO_2_) were examined at three intensities: 1) rest, 2) a submaximal intensity (respiratory exchange ratio ∼1.0), and 3) peak intensity. One-way repeated measures analysis of variance (ANOVA) models were used to determine the effects of 8- and 16-weeks of at-home HIIT on physiological data, training adherence, training compliance, and reported measures of self-efficacy. Additionally, a one-way ANOVA model was used to detect differences in training compliance in terms of achieving target heart rate zones between the first eight weeks and the second eight weeks of HIIT. When appropriate, a Bonferroni post-hoc test was used to correct for multiple comparisons and to determine where differences occurred. Additionally, a linear regression was performed to examine the relationship between the age-predicted maximal heart rate and the maximal heart rate achieved during the first graded exercise test to examine if the age-predicted maximal heart rate would serve as a good ‘target’ among this patient population. All statistics were performed using SigmaStat (Version 4, Systat, Palo Alto, CA), and *a priori* statistical significance was set as *P*<0.05. Descriptive statistics are presented as mean ± standard deviation (SD).

## RESULTS

### Graded Exercise Tests

The effects of HIIT on cardiorespiratory parameters during the graded exercise test are presented in **Table 3**. There was no effect of HIIT on resting cardiac output, heart rate, or stroke volume (*P*>0.252). However, submaximal cardiac output decreased by ∼17% following 16 weeks of at-home HIIT (*P*=0.028). There was no significant effect of HIIT on heart rate nor stroke volume at submaximal intensity (*P*>0.749, power output: 36±6 W). There was no significant effect of at-home HIIT on peak cardiac output, stroke volume, or heart rate (*P*>0.361). Additionally, VLJO_2_peak did not significantly improve with HIIT (*P*=0.064). However, peak power output improved by ∼26% following HIIT (Baseline: 70±14 W, Week 8: 82±12 W, and Week 16: 88±15 W, *P*=0.027).

**Table 3.**
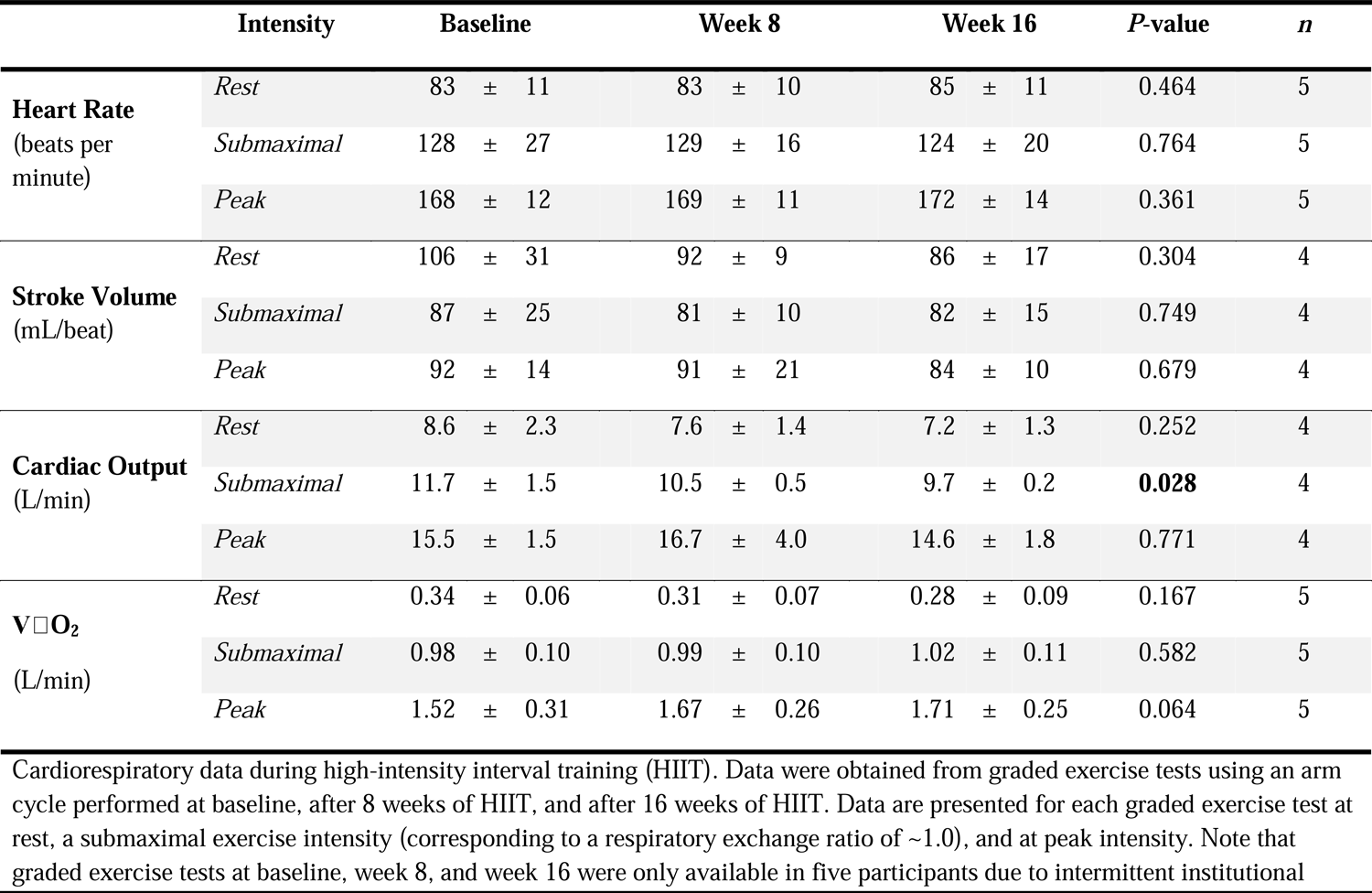

**Table 4.**
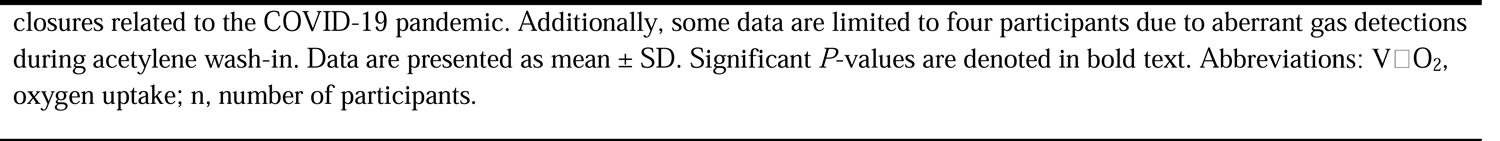

### Training Adherence and Compliance

Data regarding HIIT adherence and compliance are presented in **Table 1**. Overall adherence to the training was 87±20% (Range: 38-100%). There were no significant differences in adherence rates between the first eight weeks and the last eight weeks of at-home HIIT (*P*=0.369). Within the 24-minute HIIT sessions, participants demonstrated good compliance in achieving a heart rate corresponding to 70% HRR (80±21%, Range: 43-99%). However, participants were less compliant in achieving the target heart rate corresponding to 80% HRR during training (57±28%, Range: 9-92%).

**Figure 2** depicts the average compliance for all eight participants during the at-home HIIT sessions. Most participants demonstrated the ability to achieve a heart rate greater than 70% HRR during the latter half of the six high-intensity intervals. Conversely, the target heart rate was frequently unachieved during the two-minute bouts of recovery. During bouts of recovery, the target heart rate was only attained ∼35% of the time (Range: 1-77%). There was no effect of training on heart rate during the recovery intervals between the first eight weeks and the last eight weeks of the training program (*P*=0.982). **Figure 3** shows the composite heart rate data for all participant training sessions (average of 253 training intervals across all participants).

**Figure 2.**
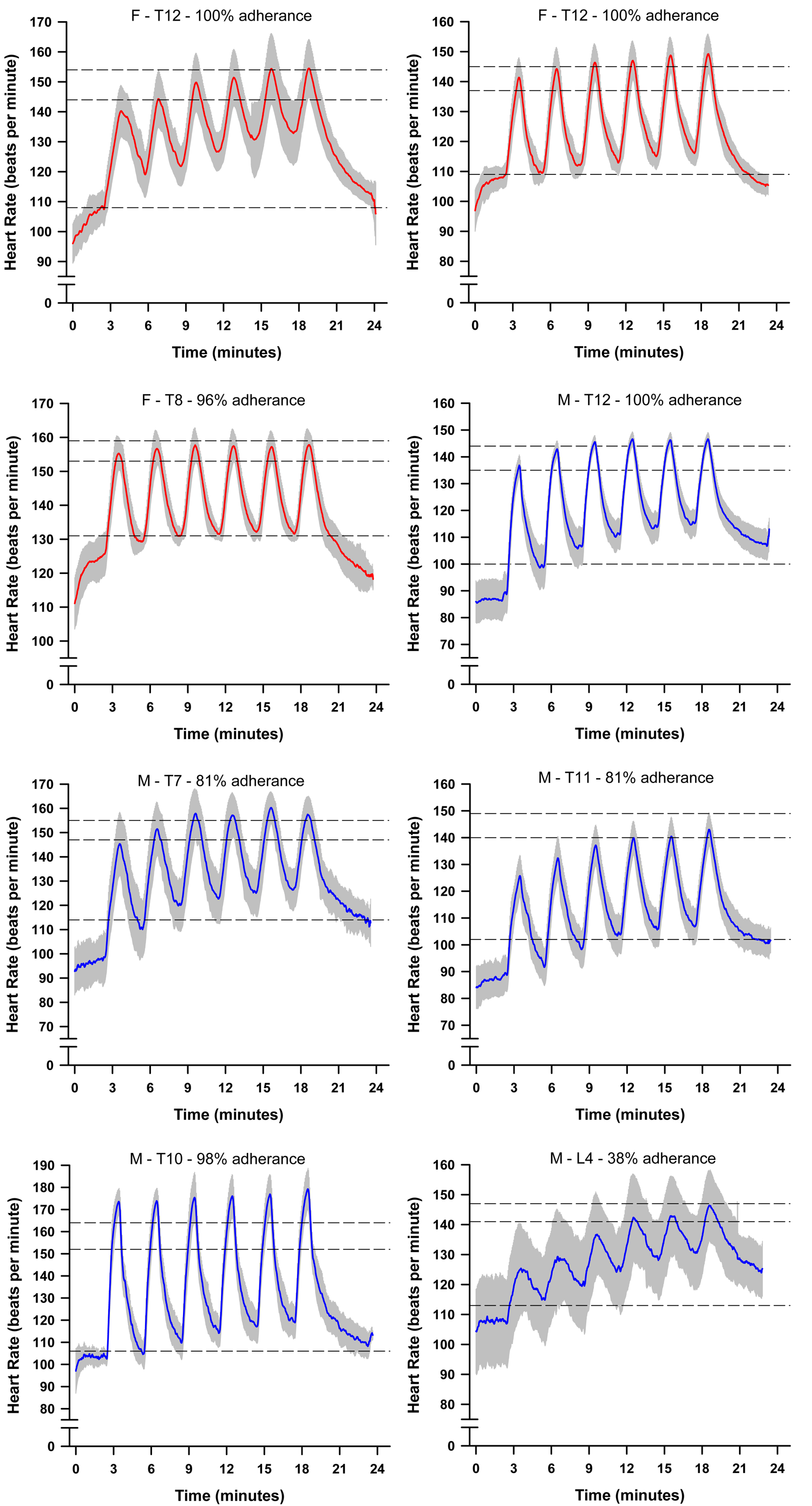
Line plots depicting individual participant heart rate data achieved during the at-home high-intensity interval training (HIIT) sessions. The solid black line depicts the average heart rate, and the grey bands depict the standard deviation. The top dashed line represents the prescribed high-intensity target of 80% heart rate reserve (HRR), the middle-dashed line represents the acceptable high-intensity threshold of 70% HRR, and bottom dashed line represents the recovery target of 30% HRR. Abbreviations: M, male; F, female; T, thoracic; HRR, heart rate reserve.

**Figure 3.**
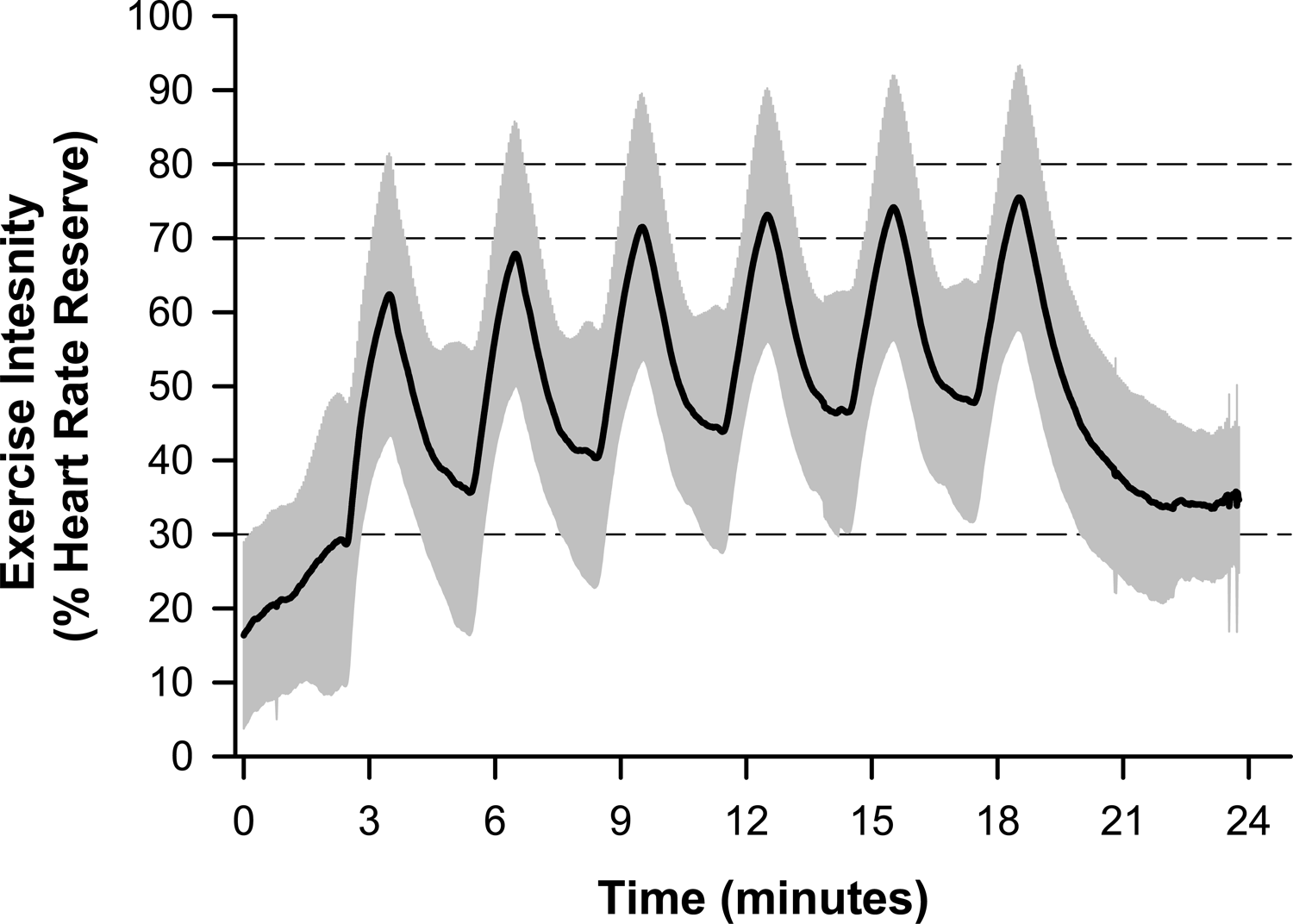
Line plot depicting the composite high-intensity interval training (HIIT) session data for all participants across all attempted workouts (*n*=338). Data is normalized to the percent of heart rate reserve (HRR). The solid black line depicts the average heart rate, and the grey bands depict the standard deviation. Dashed lines at 70 and 80% HRR represent acceptable and target HRR for each interval, respectively, and the dashed line at 30% HRR represents the recovery target.

Additionally, a linear regression demonstrated a significant relationship between the age-predicted maximal heart rate and the measured maximal heart rate during the graded exercise test (*P*=0.025, **Figure 4**).

**Figure 4:**
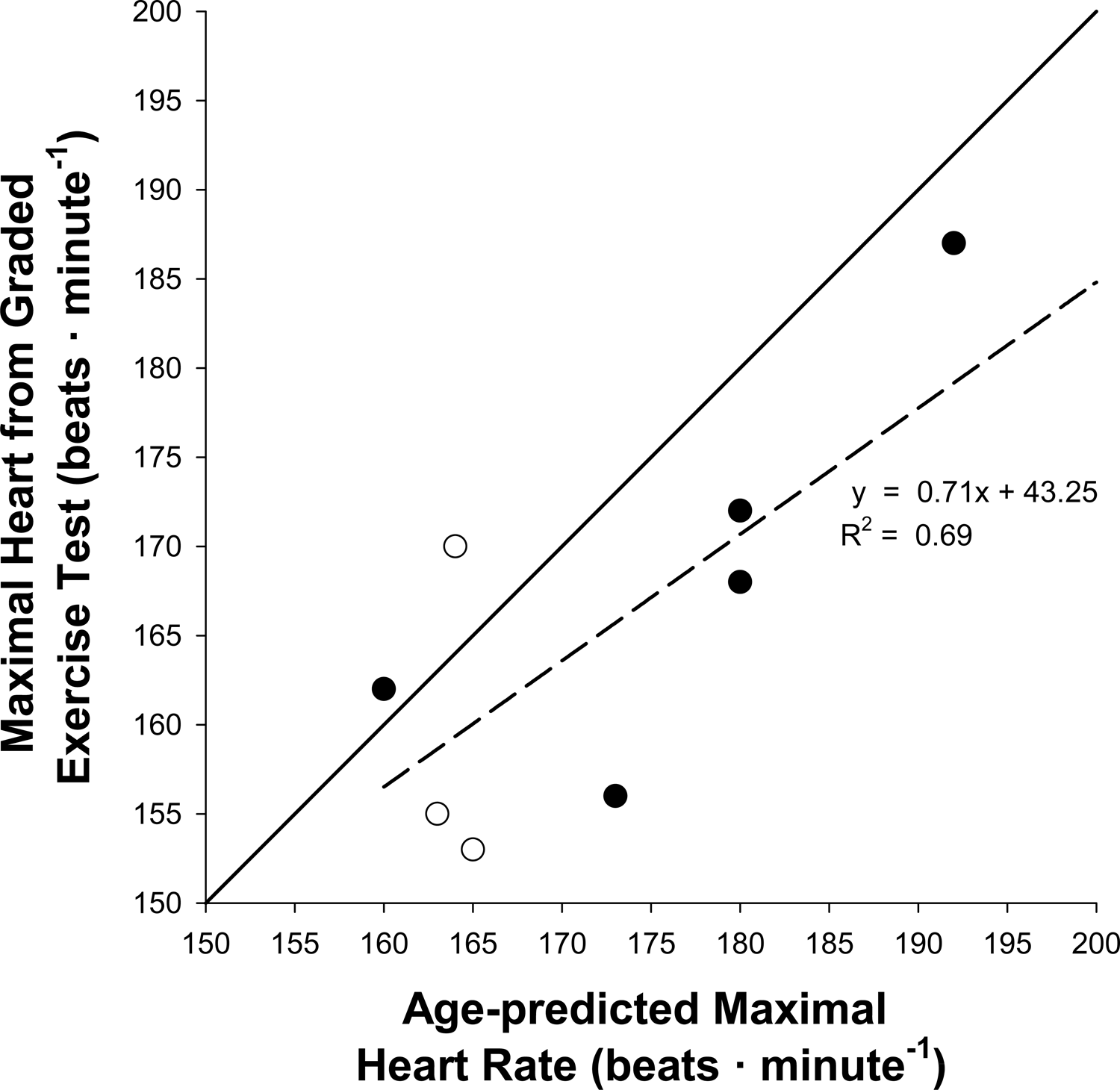
Scatter plot depicting the comparison of age-predicted maximal heart rate versus the measured maximal heart rate during a graded exercise test using arm ergometry. The solid line represents the line of identity and dashed line represents line of best fit. Solid symbols indicate male data and open symbols indicate female data.

### Participant Safety and Self-Efficacy

A total of four non-serious adverse events were reported from weekly surveys during the study. Two incidents of shoulder soreness were reported after a training session, as well as two incidents of sustained tachycardia that were deemed likely related to the intervention by the principal investigator (KLG). All incidences of tachycardia resolved within 30 minutes after the session ended, and all incidences of shoulder soreness resolved within a week. Only one participant missed a week of training due to shoulder soreness. Scores determined from the SCI ESES survey did not significantly change throughout the training period (*P*=0.497). Overall, participants reported high exercise self-efficacy scores at baseline (Total score: 3.4±3.3). However, the score in response to the statement “when confronted with a barrier to exercise I could usually find several solutions to overcome the barrier” was greater at week eight rather than week 16 (Total score: 3.6±0.5 vs 3.0±0.5, respectively, *P*=0.023). All participants were satisfied with the HIIT exercise program (**Table 2**) and agreed that they would recommend the activity to a friend, and most said they planned to continue HIIT as part of their regular exercise routine. Additionally, three participants reported a change in physical activity outside of the HIIT program. Participant four did not complete the correct survey and was lost to follow-up.

## DISCUSSION

This study aimed to assess the efficacy of a 16-week at-home high-intensity interval training (HIIT) program among individuals with spinal cord injury (SCI). Overall, participants demonstrated improvements in physical fitness, as evidenced by the elevation of peak power output (maximal work capacity) and reduced cardiac output during submaximal exercise (better exercise economy). Additionally, training adherence was high, and most participants felt positively regarding their confidence in the ability to continue regular exercise. During at-home training sessions, there was good compliance in achieving high-intensity heart rates; however, the target recovery heart rate between bouts of high-intensity was seldomly achieved due to the slow recovery kinetics of heart rate.

### Cardiorespiratory Fitness

The presented data suggest that the HIIT program improved maximal work capacity and exercise economy, as evidenced by the increase in peak power output and decrease in submaximal cardiac output, respectively. These findings are generally in good agreement with other studies examining the effects of HIIT on cardiorespiratory fitness among individuals with SCI (15–17). However, a statistically significant improvement in VLJO_2_peak was not observed among the participants following 16 weeks of HIIT. This observation may be explained by the relatively short duration of the high-intensity intervals (∼2 minutes each) used in this study, compared to the longer duration of high-intensity intervals (∼3 to 8 minutes each) used among other studies (13,22,23). Additionally, it is likely that the presented study was underpowered to detect such differences in VLJO_2_peak. Future studies warrant investigation into the dependence of improved VLJO_2_peak on the prescribed HIIT parameters (i.e., intensity and duration of intervals).

One appealing aspect of HIIT is the potential to reduce the overall exercise time per week required to elicit similar improvements in cardiometabolic health compared to continuous aerobic training. This proposed benefit may be of particular interest among participants with SCI due to the high risk of stress-induced injuries in the shoulder joint. Yet, greater durations of high-intensity intervals than investigated may be required to elicit significant improvements in VLJO_2_peak, and theoretically other cardiometabolic health factors. Therefore, there appears to be a balance in the prescribed ‘dose’ of HIIT among this population. On one hand, too little HIIT may not elicit improvements in cardiometabolic health, yet excessive HIIT may lead to stress injuries in the upper extremities that may worsen with continuous training. Future research warrants the investigation of an increased training load of HIIT on shoulder health and function to better dissociate the advantages and disadvantages of prescribed HIIT among individuals with SCI. With additional efforts, we may be able to prescribe an optimal ‘dose’ of HIIT, likely contingent on individual characteristics such as strength, baseline fitness, and fatiguability.

### Adherence

Several elements of this study may have contributed to the relatively high training adherence. First, in-home and wheelchair-accessible equipment served to remove reported barriers. A recent survey of individuals with SCI found that not having exercise equipment in the home reduced the likelihood of exercising by ∼68% (24). Additionally, the relatively short exercise durations inherent to HIIT may have also improved training adherence. Lastly, the at-home availability of the wheelchair-accessible arm cycle may have facilitated independent exercise due to enhanced availability. Independence has been noted as a motivator for consistent activity for many with SCI (9). However, it is noteworthy to highlight that high adherence rates may have also been affected by the inherent bias of the recruited participant population. The participants who were willing to be involved in this study may have a greater internal motivation to perform regular exercise compared to the general population with SCI.

### Compliance

The target heart rates during HIIT were tailored for each participant based on results from the baseline graded exercise test. Previous reports have suggested that individuals with SCI demonstrate a similar hemodynamic response during maximal exercise compared to able-bodied individuals (25). Additionally, continuous heart rate feedback provided through the smartphone application may have assisted in the achievement of the high-intensity heart rate targets. The difficulty noted with participants achieving the target recovery heart rate (∼30% HRR) may indicate that more than two minutes of recovery or a lightened recovery work rate is necessary. Altogether, these findings may help guide the future prescription of HIIT programs for individuals with SCI.

It is unclear whether adequate recovery (achieving the target recovery heart rate) influences the achievement of subsequent high-intensity bouts. Venous pooling is a known occurrence for individuals with SCI during arm cycling, which affects the hemodynamic response to exercise and may lead to an elevated heart rate between high-intensity bouts (26). Although few participants wore compression stockings during training sessions, there is limited data to suggest a significant affect upon the venous system and cardiovascular responses (27). In light of the presented findings, future work is needed to examine the changes in heart rate during HIIT training among individuals with SCI to determine the optimal duration of high-intensity and recovery intervals necessary to achieve the prescribed HRR targets. Additionally, these findings demonstrated a positive association between the maximal heart rate achieved during the baseline graded exercise test to the age-predicted maximal heart rate. Therefore, we suggest that age-predicted heart rate may be a useful target when prescribing exercise for this specific SCI population (injury below T6).

### Participant Safety and Self-Efficacy

Infrequent incidences of adverse events were reported during the at-home HIIT program. However, indications of tachycardia were noted by primary care providers prior to study involvement, with no notable concern about participating in a regular exercise program. Reports of shoulder pain were addressed by advising rest and reduced resistance during the HIIT sessions. Additionally, some investigators have reported improvements in shoulder health among individuals with SCI following upper extremity exercise. For instance, Graham *et al.* noted an improvement in strength scores following arm cycle training (14). From these results, the authors suggest that only 40 minutes of HIIT per week was sufficient to improve upper extremity strength in the chest press and lateral pull-down. Although the upper extremity adverse events were minor in our study, future studies focusing on upper extremity exercise should address shoulder health in screening and throughout the study by denoting any alterations in shoulder pain, range of motion, and strength.

Within this study, participants reported moderate confidence in exercise participation at baseline, and this confidence persisted throughout the study. Given their willingness to participate in a 16-week exercise study, it is possible that the recruited participants may have greater confidence in exercise relative to the general SCI population. Additionally, participants reported a lower barrier to exercise after eight weeks of HIIT than at the end of the study. The lower confidence in overcoming these barriers at the end of the study could potentially be due to the need to return the arm cycle and other equipment to the study staff following termination of the HIIT program.

Satisfaction survey data indicated that most participants enjoyed this at-home HIIT program. Similarly, HIIT has previously been demonstrated to be more enjoyable than moderate-intensity exercise in the SCI population (28,29). Some have suggested that the reported enjoyment may be associated with the frequent recovery bouts breaking up the monotony of the workout, the constant change in activity, and the feeling of accomplishment achieved through more intense work (28). Despite COVID-19 pandemic-related restrictions at our institution during the study (March 2020 through July 2021), participants continued to perform the HIIT program at home.

## Conclusion

This pilot study suggests that a 16-week at-home HIIT program was easily implemented for individuals with SCI below the sixth thoracic vertebrae. Overall, participants achieved high adherence and compliance, infrequent adverse events, a high reported self-efficacy, and improvements in exercise economy and maximal work capacity. From these observations, we propose that HIIT may provide an enjoyable long-term aerobic exercise program for individuals with SCI. Future studies warrant rigorous examination in relation to sedentary controls and other training programs.

## Data Availability

All data produced in the present study are available upon reasonable request to the authors

## Acknowledgements

We would like to thank Andrew Miller, Technical Specialist Coordinator within the Clinical Research and Trials Unit at Mayo Clinic for conducting maximal exercise testing for all participants. We would like to thank Tyson Scrabeck and Julie Block for working with participants during the COVID-19 pandemic to navigate clinical changes. We would like to thank the Mayo Clinic Media Services Department for assisting in editing the workout recordings. We would like to sincerely thank our participants for their dedication to the 16-week project. The authors declare that the results of the study are presented clearly, honestly, and without fabrication, falsification, or inappropriate data manipulation. The results of the present study do not constitute endorsement by ACSM.

## Author Contributions

KLG, MBL, and DDV initiated the project. MBL, KLW, CCW and DDV, communicated directly with patients and carried out patient education and testing. MBL and KLW drafted the manuscript and incorporated all author edits. CCW and KLW contributed to writing the report, extracting, analyzing, and interpretation of data, OHM assisted in data analysis. DDV, MLG, MGV, ERL, OHM, MJJ, LAB, KDZ and KLG contributed edits to the manuscript. KLG supervised all aspects of this project and provided approval for the publication and content.

## Funding

This work was funded, in part, by the Mayo Clinic Rehabilitation Medicine Research Center, which received benefactor funding. KLG received funding from the Mayo Clinic Center for Clinical and Translational Science Small Grants Program Fund. CCW was supported by a National Institutes of Health Training Grant (T32-DK-007352-39). KLW was supported by a National Institutes of Health Training Grant (T32-HL105355-10) and the Mayo Clinic Graduate School of Biomedical Sciences.

## Ethical Approval

All procedures have been approved and performed in accordance with the Declaration of Helsinki and by the Mayo Clinic Institutional Review Board #18-004972. We certify that all applicable institutional and governmental regulations concerning the ethical use of human volunteers were followed during the course of this research.

## Competing Interests

The authors declare no competing interests in relation to the work described.

## Data Availability

The datasets generated during and/or analyzed during the current study are available from the corresponding author upon reasonable request.

## Notes

### Competing Interest Statement

The authors have declared no competing interest.

### Clinical Trial

NCT04378218

### Author Declarations

All experimental procedures were approved by the Mayo Clinic Institutional Review Board (IRB# 18-004972, Clinical Trials Identifier: NCT04378218). Experimental procedures were performed in accordance with the ethical standards set by the Declaration of Helsinki, excluding registration in a public database. Participants provided written informed consent prior to enrollment.

