## Supplemental Table 1 for "At-Home High-Intensity Interval Training for Individuals with Paraplegia Following Spinal Cord Injury: *A Pilot Study*"

**Supplementary Table 1.**

| **Question** |  |
| --- | --- |
| Q1 | In the past week, how many times did you complete a HIIT workout on the arm-cycle? |
| Q2 | During the workouts, were you able to reach your target heart rate during the HIIT intervals? |
| Q3 | In the past week, did you experience any pain during or after the HIIT workout that limited the use of your arms for an extended period of time? |
| Q4 | Do you have any injuries or concerns to report that will prevent you from completing future HIIT sessions? |
| Q5 | Do you have any injuries or concerns to report that will cause you to leave the study early? |
| Weekly survey to determine participant-reported levels of safety. Abbreviations: Q, question. | |
